# Characteristics and Long-term Mortality in Patients with ST-Segment Elevation Myocardial Infarction with Non-Obstructive Coronary Arteries (STE-MINOCA): A High Risk Cohort

**DOI:** 10.1101/2023.02.05.23285502

**Authors:** Odayme Quesada, Mehmet Yildiz, Timothy D. Henry, Brynn K. Okeson, Jenny Chambers, Ananya Shah, Larissa Stanberry, Lucas Volpenhein, Dalia Aziz, Rebekah Lantz, Cassady Palmer, Justin Ugwu, Muhammad J. Ahsan, Ross F. Garberich, Heather S. Rohm, Frank V. Aguirre, Santiago Garcia, Scott W. Sharkey

## Abstract

**Background:** The prognosis of ST-segment elevation myocardial infarction with non-obstructive coronaries (STE-MINOCA) is largely unknown.

**Methods:** The objective of this study is to evaluate the prevalence, characteristics, and 5-year mortality of patients with STE-MINOCA compared to STEMI with coronary artery obstruction (STEMI-Obstruction) using a multicenter cohort of consecutive STEMI patients at 3 regional Midwest STEMI programs from 2003 to 2020. STE-MINOCA was defined based on (1) coronary stenosis < 60% by visual estimation, (2) ischemia with elevated troponin, and (3) no alternative diagnosis. STE-MINOCA was further classified based on American Heart Association (AHA) definition as AHA STE-MINOCA and AHA STE-MINOCA Mimicker.

**Results:** 8,566 STEMI patients, including 420 (4.9%) STE-MINOCA (26.9% AHA STE-MINOCA and 73.1% AHA STE-MINOCA Mimicker) were followed for a median of 7.1 years. Compared to STEMI-Obstruction, STE-MINOCA were younger, more often female, had fewer cardiovascular risk factors, and were less likely to be discharged on cardiac medications. At five years, mortality was higher in STE-MINOCA compared with STEMI-Obstruction (18% vs. 15%, p=0.033). In propensity score-matched analysis, STE-MINOCA had a 1.4-fold (95% CI: 1.04-1.89, p=0.028) higher risk of 5-year all-cause mortality compared with STEMI-Obstruction. Furthermore, 5-year mortality risk was significantly higher in AHA STE-MINOCA Mimicker (19% vs. 15%, p=0.043) but similar in AHA STE-MINOCA (17% vs. 15%, p=0.42) compared with STEMI-Obstruction.

**Conclusions:** In this large multicenter STEMI cohort, nearly 5% of patients presented with STE-MINOCA. At five years, mortality approached 20% among patients with STE-MINOCA. Despite the lower risk profile, STE-MINOCA patients were at 40% higher risk of 5-year all-cause mortality compared with STEMI-Obstruction. Additionally, 5-year all-cause mortality risk was higher in AHA STE-MINOCA Mimicker but similar in AHA STE-MINOCA compared to STEMI-Obstruction.

## INTRODUCTION

Myocardial infarction with non-obstructive coronary arteries (MINOCA) occurs in 5-15% of all acute myocardial infarction (AMI) cases referred for coronary angiography (1–3). MINOCA consists of a heterogeneous group of etiologies characterized by ischemia and the absence of obstructive coronary artery disease (CAD) (4). Among ST-segment elevation MI (STEMI) presentations, 4.9-6.6% are STE-MINOCA (2,5).

MINOCA has historically been considered a lower-risk AMI phenotype. However, several recent studies have documented worse outcomes in MINOCA than in a general population without known cardiovascular disease (CVD) (6–8). When MINOCA cases are compared to those with AMI and obstructive CAD, many studies have reported worse outcomes in obstructive CAD (1,2,8–10), while others have noted similar or higher mortality risk in MINOCA (11–14). These disparate findings are predominantly due to: 1) inconsistencies in the use of an elevated troponin to define MINOCA; 2) MINOCA definitions based on the 2019 American Heart Association (AHA) (15) versus 2016 European Society of Cardiology (ESC) (16) statements; 3) the comparator group (general population without known CVD versus patients with obstructive CAD); and 4) the electrocardiographic type of AMI at presentation (i.e., NSTEMI (non-ST-segment elevation myocardial infarction) versus STEMI).

MINOCA studies to date are predominantly composed of patients with NSTEMI; reports of STE-MINOCA are infrequent (5,17–20). Therefore, the characteristics and prognosis of patients with STE-MINOCA are largely unknown. To further inform this area of uncertainty, we examined the Midwest STEMI Consortium database to evaluate the prevalence, characteristics, and 5-year mortality of STE-MINOCA patients compared to STEMI with coronary artery obstruction (STEMI-Obstruction). We also examined these differences in STEMI-Obstruction compared to AHA-defined STE-MINOCA and STE-MINOCA Mimicker.

## METHODS

### Study Population

The Midwest STEMI Consortium is composed of high-volume, tertiary, regional STEMI centers, including The Christ Hospital in Cincinnati, OH; Minneapolis Heart Institute in Minneapolis, MN; Prairie Heart Institute in Springfield, IL; and Iowa Heart Center in Des Moines, IO (21). All consecutive patients from March 2003 to December 2020 who presented with ST-segment elevation ≥1mm in at least two contiguous leads or new (or presumably new) left bundle branch block within 24 hours of symptom onset were prospectively included into the consortium database. These regional tertiary centers use standardized STEMI protocols in their region and serve as the referral center for over 100 non-percutaneous coronary intervention (PCI) hospitals within a spectrum of urban and rural communities. The design of the Midwest STEMI consortium has been described in detail previously (21). The study protocol, data sharing agreements, and relevant information have been approved by Institutional Review Boards (IRB) in each study center. The American College of Cardiology Accreditation Services initially funded the Midwest STEMI Consortium.

The study flow diagram is shown in **Figure 1**. Patients with prior coronary artery bypass graft surgery (CABG) and those who received fibrinolytic therapy on presentation were excluded from this study. Data from Iowa Heart Center was not included in this study, given the inability to provide 5-year mortality data.

**Figure 1.**
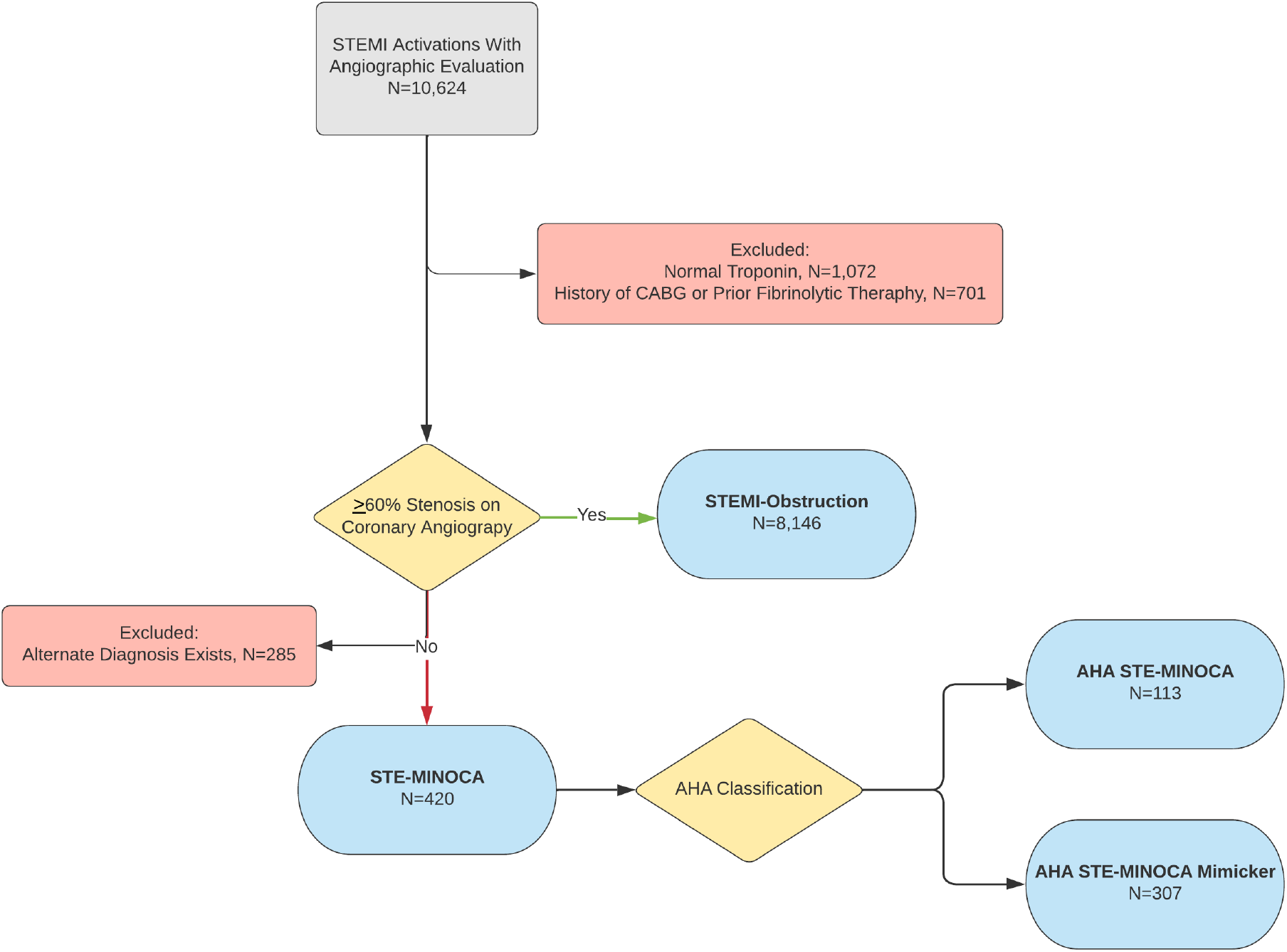
Study Flow Chart. The study population was derived from a multi-center, prospective Midwest STEMI consortium registry.

STE-MINOCA was defined as (1) ST-segment elevation ≥1mm in at least two contiguous leads or new (or presumably new) left bundle branch block with symptoms of myocardial ischemia, (2) absence of stenosis ≥60% in any major epicardial vessels during invasive coronary angiography by visual estimation, (3) evidence of ischemia defined as at least one troponin measurement >99^th^ percentile, and (4) absence of an alternative diagnosis in the index presentation. SCAD cases with an obstructive culprit lesion were classified as STEMI-Obstruction, and those without obstructive lesions were classified as STE-MINOCA. As per the ESC statement, takotsubo cardiomyopathy, myocarditis, and non-ischemic cardiomyopathy were included in the MINOCA cohort (16).

In a secondary analysis, we further categorized the STE-MINOCA based on the 2019 AHA statement as either AHA STE-MINOCA (confirmed or clinically suspected for coronary artery plaque disruption, epicardial coronary spasm, coronary embolism/thrombosis) or AHA STE-MINOCA Mimicker (confirmed or clinically suspected takotsubo cardiomyopathy, myocarditis, and non-ischemic cardiomyopathy) (15).

### Data Collection and Follow-up

At the time of STEMI, trained research assistants prospectively collected all relevant clinical data, including demographic and clinical characteristics, angiographic details, laboratory markers, and discharge medications by reviewing electronic medical records. Left ventricular ejection fraction was collected from an echocardiogram, performed pre- or just after the coronary angiography during the index hospitalization. The National Death Index (NDI), a reliable tool to identify dead subjects, and electronic medical record review were used to determine mortality at the 5-year follow-up.

Each AHA STE-MINOCA case was retrospectively classified for this study using medical records and a review of coronary angiographic images by the principal investigator at each site. Coronary intravascular imaging and cardiac magnetic resonance (CMR) imaging results were not uniformly utilized, but, when available, were used for diagnosis. In cases of disagreement, the case was presented and discussed among two principal investigators to arrive at the final classification. In only one study site, Minneapolis Heart Institute, CMR was routinely recommended in the workup of MINOCA cases, and the CMR findings were investigated in more detail in the secondary analysis.

### Statistical Analyses

Categorical variables are presented as numbers and relative frequencies (percentages) and continuous variables as mean ± SD or median (IQR). Demographics, clinic characteristics, medications at discharge, and outcomes were compared between STE-MINOCA and STEMI-Obstruction. Categorical variables were compared using the Chi-square test or Fischer’s exact test and continuous variables using an independent sample t-test or Wilcoxon rank-sum test. The primary endpoint of the study was 5-year survival. Survival of patients in the STEMI-Obstruction and STE-MINOCA from the time of STEMI presentation to 5 years was estimated using the Kaplan-Meier method and compared using the log-rank test.

A propensity score-matched sample was created using nearest-neighbor matching. Propensity scores were estimated using logistic regression of the STEMI group on the following variables: age, sex, hypertension, diabetes, dyslipidemia, PCI history, site, and year of admission. Matching variables were selected out of known baseline risk factors based on current literature and availability in the data; therefore, 8,288 of 8,566 (97%) patients with complete data were included in the matching procedure. Matching was performed without replacement based on the logit of the propensity score with a ratio of 1:5 and using a caliper width of 0.2 standardized differences. One STE-MINOCA patient was unmatched, resulting in a matched sample of 2,367 patients. Hazard ratios were estimated from the matched sample via a Cox regression model adjusted for the matching variables and PCI history. The interaction term between group (STEMI-Obstruction or STE-MINOCA) and PCI history was found insignificant and removed from the model. A second Cox regression model was adjusted for the same covariates as the first, with the addition of troponin. Age and log-transformed troponin were modeled with splines due to nonlinearity. The model violated the proportional hazards assumption and was therefore stratified by sex. The site was included in the model as a random effect. R version 4.1.1 and RStudio (R Core Team 2021) were used for all statistical analyses.

In the secondary analysis, demographics, clinic characteristics, medications at discharge, and outcomes were compared between STEMI-Obstruction and AHA STE-MINOCA and AHA STE-MINOCA Mimicker. The 5-year survival of patients in the STEMI-Obstruction, AHA STE -MINOCA, and AHA STE-MINOCA Mimicker groups from the time of STEMI presentation to 5-year was estimated using the Kaplan-Meier method and compared using the log-rank test.

## RESULTS

### Demographic and Clinical Characteristics in STE-MINOCA versus STEMI-Obstruction

We identified a total of 10,624 consecutive STEMI activations from 2003 to 2020. Of these activations, 2,058 (19%) were excluded for the following reasons: Normal troponin levels (n=1,072), history of CABG or prior fibrinolytic therapy (n=701), or alternate diagnosis (n=285), including demand ischemia such as sepsis, pulmonary embolism, relook angiograms on patients with recent interventions leading to 8,566 included in the final analysis (**Figure 1**).

Among 8,566 STEMI patients, 420 (4.9%) were STE-MINOCA, and 8,146 (95.1%) were STEMI-Obstruction. The frequency of STE-MINOCA was not significantly different among the three sites (4.9% Minneapolis Heart Institue, 5.3% Prairie Heart Institute, and 4% The Christ Hospital; p=0.2). Furthermore, when the cohort was divided by decade, the frequency of STE-MINOCA was not significantly different between 2003-2012 and 2013 to 2020 (X^2^=2.80, p=0.094). Distribution of the frequency to STE-MINOCA during the enrollment period by year is illustrated in **Supplemental Figure 1**. Compared to STEMI-Obstruction, patients with STE-MINOCA were significantly younger, more frequently female, and had less cardiovascular risk factors (**Table 1**).

**Table 1.** Demographics, Clinical and Angiographic Characteristics in STE-MINOCA and STEMI-Obstruction

|  | STE-MINOCA<br>N=420 | STEMI-Obstruction<br>N=8,146 | p Value |
| --- | --- | --- | --- |
| **Demographics** |  |  |  |
| Age, y | 59 ± 18 | 62 ± 13 | 0.002 |
| Female | 222 (52.9) | 2,386 (29.3) | <0.001 |
| **Comorbidities** |  |  |  |
| Hypertension | 212 (52) | 4,922 (61) | <0.001 |
| Dyslipidemia | 169 (42) | 4,305 (54) | <0.001 |
| Diabetes | 62 (15) | 1,644 (20) | 0.012 |
| Smoking history | 88 (24) | 5,050 (64) | <0.001 |
| Family history of CAD | 135 (40) | 2,931 (48) | 0.003 |
| Previous stroke | 4 (1.6) | 218 (4.3) | 0.039 |
| Previous PCI | 46 (11) | 1,643 (20) | <0.001 |
| **Clinical characteristics** |  |  |  |
| SBP, mm Hg | 137 ± 28 | 140 ± 32 | 0.13 |
| DBP, mm Hg | 83 ± 19 | 84 ± 21 | 0.32 |
| Heart rate, bpm | 86 (73, 103) | 76 (64, 90) | <0.001 |
| Out-of-hospital cardiac arrest | 20 (8.1) | 670 (9.0) | 0.61 |
| Cardiogenic shock (pre-PCI) | 15 (5.4) | 580 (7.8) | 0.14 |
| Killip class |  |  | 0.16 |
| I/II | 334 (93) | 6,801 (91) |  |
| III/IV | 24 (6.7) | 662 (8.9) |  |
| LVEF, % | 45 (30, 55) | 50 (40, 55) | <0.001 |
| **Laboratory markers** |  |  |  |
| Troponin, ng/ml | 1.42 (0.31, 5.07) | 8.38 (1.90, 39.30) | <0.001 |
| Baseline creatinine, mg/dl | 0.90 (0.77, 1.10) | 1.00 (0.84, 1.20) | <0.001 |
| HDL, mg/dl | 42 (33, 51) | 37 (31, 44) | <0.001 |
| LDL, mg/dl | 90 (66, 110) | 100 (76, 125) | <0.001 |
| Triglycerides, mg/dl | 105 (78, 135) | 124 (87, 175) | <0.001 |
| Total cholesterol, mg/dl | 154 (127, 184) | 166 (138, 195) | <0.001 |
| **Angiographic characteristics** |  |  |  |
| **Number of diseased vessels** |  |  | - |
| 0 | 221 | 0 |  |
| 1 | 0 | 2,776 |  |
| 2 or more | 0 | 2,760 |  |
| **Culprit vessel** |  |  | - |
| Left main | 0 | 77 (0.9) |  |
| LAD | 0 | 3,177 (39) |  |
| LCx | 0 | 1,182 (15) |  |
| RCA | 0 | 3,499 (43) |  |
| Other (Branches) | 0 | 104 (1.3) |  |
| Multiple | 0 | 107 (1.3) |  |
| None | 420 (100) | 0 (0) |  |
| **TIMI flow (pre-PCI)** |  |  | - |
| 0/1 | 0 | 5,087 |  |
| 2/3 | 158 | 2,876 |  |
| **TIMI flow (post-PCI)** |  |  | - |
| 0/1 | 0 | 179 |  |
| 2/3 | 152 | 7,778 |  |

Values are n (%), mean ± SD, and median (IQR)
CAD = coronary artery disease; DBP = diastolic blood pressure; SBP = systolic blood pressure; PCI = percutaneous coronary intervention; LVEF = left ventricular ejection fraction; HDL = high-density lipoprotein; LAD = left anterior descending; LCx = left circumflex; LDL = low-density lipoprotein; and RCA = right coronary artery

The frequency of out-of-hospital cardiac arrest and cardiogenic shock (pre-PCI) were not significantly different between the two groups (**Table 1**). However, patients with STE-MINOCA had significantly lower left ventricular ejection fraction and lower troponin levels than those with STEMI-Obstruction. In addition, total cholesterol, LDL cholesterol, and triglycerides were significantly lower in STE-MINOCA. Compared to those with STEMI-Obstruction, patients with STE-MINOCA were less likely to be discharged on aspirin, P2Y12 inhibitors, beta-blockers, renin-angiotensin system blockers [angiotensin-converting-enzyme inhibitor (ACEI)/angiotensin receptor blocker (ARB)], and statins (**Table 2**).

**Table 2.**
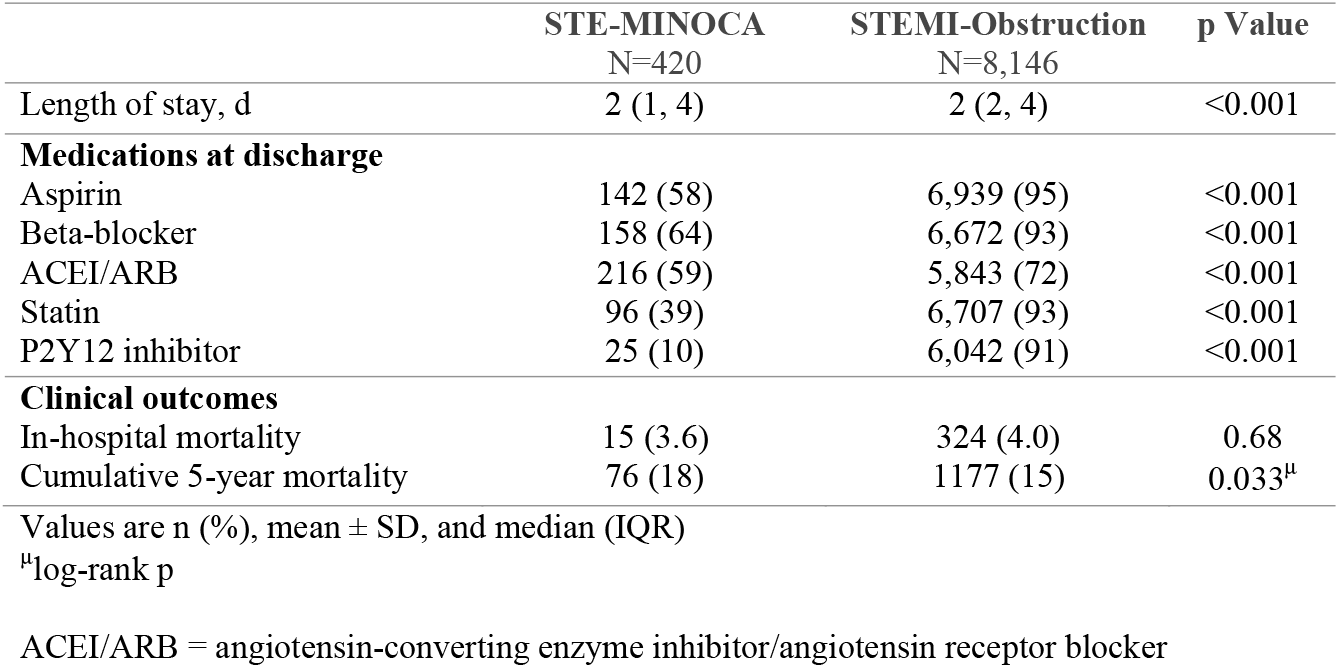
Discharge Medications and Outcomes in STE-MINOCA and STEMI-Obstruction

### In-Hospital and 5-Year Mortality

In-hospital mortality risk was similar between STE-MINOCA and STEMI-Obstruction (**Table 2**). Median follow-up for mortality was 7.1 years. The crude estimated risk of all-cause mortality within 5 years in STE-MINOCA was 20% (95% CI: 15.8-23.9%). Overall 5-year survival probability was lower in STE-MINOCA compared with STEMI-Obstruction (log-rank p=0.033) **(Figure 2)**. Covariate balance in unmatched and matched analysis is demonstrated in **Supplemental Figure 2**. In a propensity score-matched analysis, the higher risk of 5-year all-cause mortality in STE-MINOCA compared to STEMI-Obstruction was insignificant (HR 1.19, 95% CI: 0.91-1.57, p=0.21). However, when troponin was adjusted in the Cox regression model, STE-MINOCA had a 40% higher risk of 5-year all-cause mortality than STEMI-Obstruction (HR 1.40, 95% CI: 1.04-1.89, p=0.028) **(Table 3)**.

**Table 3.**
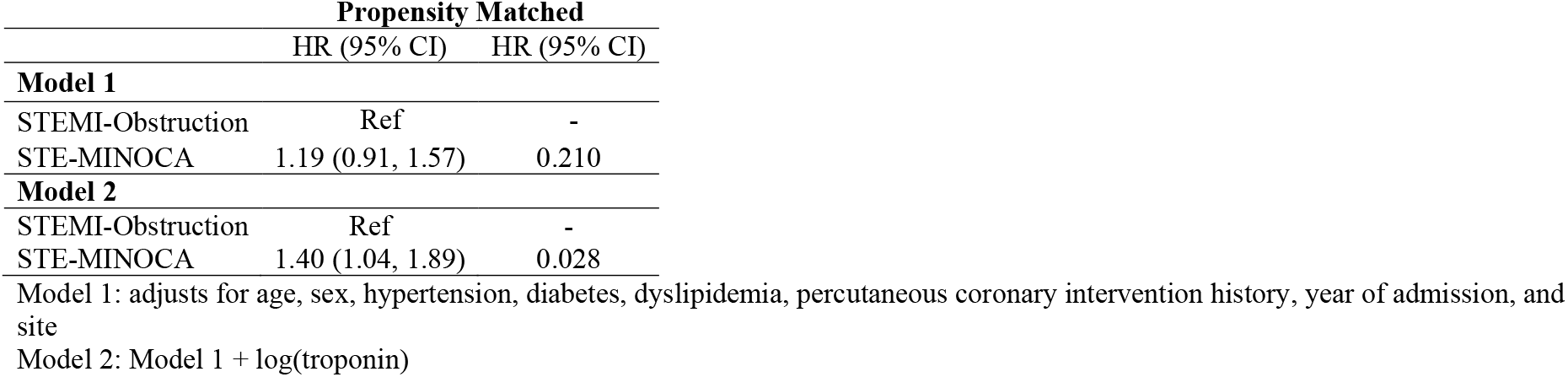
Five-Year All-Cause Mortality Risk In STE-MINOCA compared to STEMI-Obstruction

**Figure 2.**
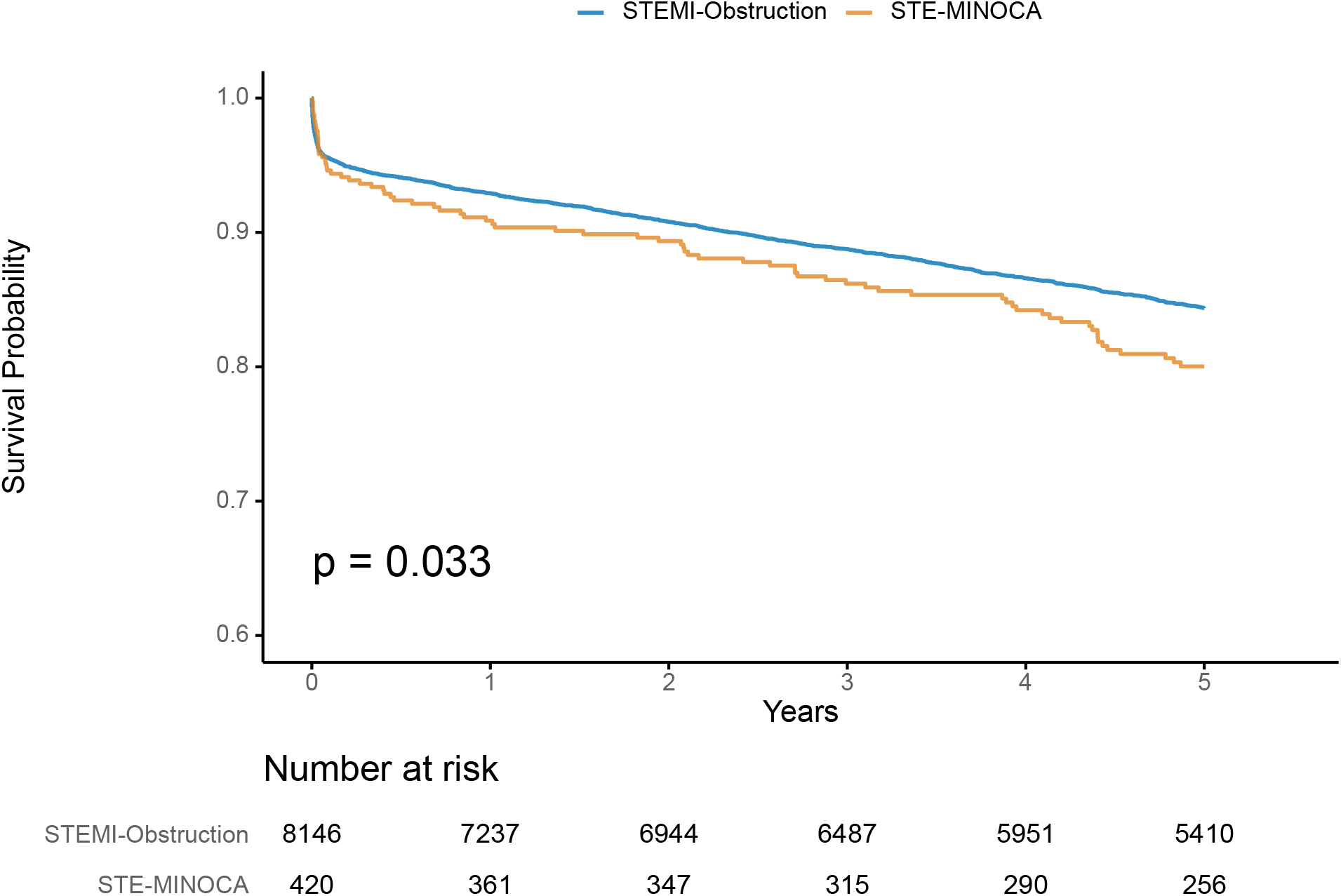
5-Year Survival Probability in STE-MINOCA in Comparison with STEMI-Obstruction. Kaplan-Meier curves with 5-year survival probability in STE-MINOCA and STEMI-Obstruction.

### Secondary Analysis: AHA STE-MINOCA and AHA STE-MINOCA Mimicker

Among the 420 patients classified as STE-MINOCA, 113 (26.9%) were further classified as AHA STE-MINOCA and 307 (73.1%) as AHA STE-MINOCA Mimicker (**Supplemental Table 1**). Among MINOCA-Mimicker, 160 (52%) had takotsubo cardiomyopathy, 108 (35%) myocarditis, and 39 (13%) non-ischemic cardiomyopathy. Compared with STEMI-Obstruction, AHA STE-MINOCA and AHA STE-MINOCA Mimicker were significantly younger and more often female. In addition, AHA STE-MINOCA Mimicker was less likely to have cardiovascular risk factors compared to STEMI-Obstruction.

Compared with STEMI-Obstruction, the Killip class and frequency of out-of-hospital cardiac arrest were similar among AHA STE-MINOCA and AHA STE-MINOCA Mimicker (**Supplemental Table 1**). Compared to STEMI-Obstruction, the left ventricular ejection fraction was significantly lower in AHA STE-MINOCA Mimicker and higher in AHA STE-MINOCA. Troponin levels were significantly higher in those with STEMI-Obstruction than in AHA STE-MINOCA and AHA STE-MINOCA Mimicker. Compared with STEMI-Obstruction, AHA STE-MINOCA and AHA STE-MINOCA Mimicker were less frequently discharged on aspirin, P2Y12 inhibitors, beta-blockers, ACEI/ARB, and statins (**Supplemental Table 2**).

At Minneapolis Heart Institute, CMR was performed in 27 (73%) of patients with AHA STE-MINOCA with the following findings: evidence of infarction 20 (74%), regional wall motion abnormality without evidence of infarction 4 (11%), and no evidence of infarct or wall motion abnormality 3 (8.1%). Of note, among the 10 AHA STE-MINOCA cases where a CMR was not performed, there was a presumed diagnosis for the cause of the event based on the coronary angiography (such as coronary embolism, epicardial coronary spasm, or SCAD). At Minneapolis Heart Institute, the number of unknown cause for AHA STE-MINOCA was significantly lower (11%) compared to The Christ Hospital (56%) and Prairie Heart Institute (43%), where CMR was not utilized routinely (p<0.001). In addition, CMR was performed in 66 (38.7%) of AHA STE-MINOCA Mimicker patients with the following findings: Takotsubo Syndrome 36 (54.5%), myocarditis 27 (40.9%), and non-ischemic cardiomyopathy 3 (4.5%).

The risk of in-hospital mortality was similar between STEMI-Obstruction and both AHA STE-MINOCA and AHA STE-MINOCA Mimicker (**Supplemental Table 2**). Cumulative 5-year mortality risk was similar between STEMI-Obstruction and AHA STE-MINOCA **(Figure 3A)**, whereas AHA STE-MINOCA Mimicker had significantly higher 5-year mortality risk compared with STEMI-Obstruction (19% vs. 15%, p=0.043) **(Figure 3B)**.

**Figure 3.**
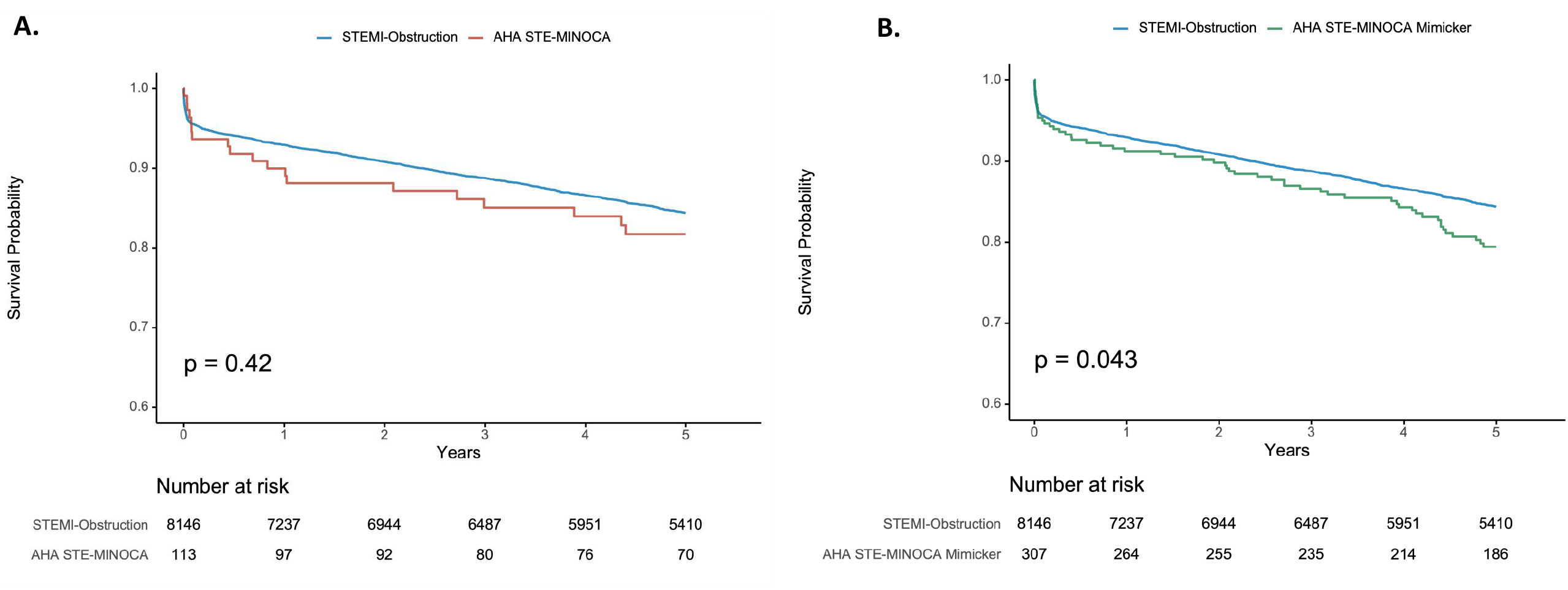
5-year Survival Probability in STEMI-Obstruction in Comparison to AHA STE-MINOCA [A] and STE-MINOCA Mimicker [B]. Kaplan-Meier curves with 5-year survival probability in STEMI-Obstruction compared to AHA STE-MINOCA [A] and STE-MINOCA Mimicker [B]

## DISCUSSION

This multicenter cohort of 8,566 consecutive STEMI patients, gathered over 20 years, identified over 400 patients with STE-MINOCA, representing the largest report to date. Several observations advance our understanding of this intriguing entity: 1) Despite the lower traditional cardiovascular risk profile, STE-MINOCA patients were at 40% higher risk of 5-year all-cause mortality compared to STEMI-Obstruction; 2) STE-MINOCA patients had a significantly lower frequency of cardiac medications prescribed at discharge; 3) when classified based on AHA statement, compared with STEMI-Obstruction 5-year all-cause mortality was higher in AHA STE-MINOCA Mimicker but similar in AHA STE-MINOCA; and 4) in some STE-MINOCA cases, CMR may provide evidence for an underlying mechanism.

The prevalence of STE-MINOCA in our study (4.9%) is similar to that reported in other published series: 6.6% in a 2021 meta-analysis of 806,851 consecutive patients with AMI (2) and 4.4% in a 2019 study by Gue et al. (5). In our study, patients with STE-MINOCA were younger, more commonly women, and had fewer traditional cardiovascular risk factors, including hypertension, dyslipidemia, diabetes, smoking, and family history of CAD, compared to STEMI-Obstruction, consistent with prior studies (17,18). STE-MINOCA also had lower peak troponin levels compared to STE-Obstruction, yet paradoxically lower ejection fraction was likely driven by AHA STE-MINOCA Mimicker. In our study, the all-cause mortality risk at 5 years for STE-MINOCA was 20%. Other reports of STE-MINOCA mortality involve shorter follow-up duration; Gue et al. reported 4.5% of all-cause mortality risk during 1-year follow-up (5), and Andersson et al. reported 8% estimated risk of mortality in a single Denmark center study, involving 4,793 consecutive STEMI patients, followed for a median of 2.6 years (18).

Among MINOCA patients, STE-MINOCA has worse outcomes than NSTE-MINOCA, and the presence of STE has been reported to be a predictor of major adverse cardiovascular events (MACE), heart failure, and mortality (19,20,22). To our knowledge, only two previous studies have compared outcomes of STE-MINOCA with STEMI-Obstruction, with conflicting findings (17,18). In a post hoc analysis of the HORIZONS-AMI trial, among 3,602 STEMI patients, 127 (3.5%) had no significant CAD at 3-year follow-up and MACE risk was lower in patients without significant CAD than in those with obstructive CAD (mortality risk was not reported) (17). The lower MACE risk in the post-hoc analysis was likely influenced by the fact that nearly 50% of patients did not have elevated cardiac biomarkers, therefore not meeting the contemporary MINOCA definition. In contrast, all patients in our cohort had elevated troponin levels. Also, the HORIZONS-AMI post hoc analysis likely represents a lower-risk MINOCA group (coronary stenosis defined as less than 30%) compared to our cohort, in which the threshold stenosis was less than 60%. Consistent with our findings, Andersson et al. reported a 2.44-fold higher risk of death in patients with normal coronary arteries compared to obstructive CAD (18).

The magnitude of troponin elevation is directly associated with mortality in patients with acute myocardial infarction (23). In our study, the peak troponin level was significantly lower in patients with STE-MINOCA compared with STEMI-Obstruction. Yet, when the mortality analysis was adjusted for peak troponin levels, patients with STE-MINOCA had significantly higher long-term mortality risk than those with STEMI-Obstruction, an anomaly that requires further investigation.

MINOCA poses a dilemma for clinicians since several etiologies and pathologic mechanisms underlie this enigmatic syndrome. Given the discordance between the AHA and ESC position statements regarding the inclusion or exclusion of takotsubo cardiomyopathy, myocarditis, and non-ischemic cardiomyopathy in the MINOCA definition (15,16), we separated our STE-MINOCA cohort into AHA STE-MINOCA and AHA STE-MINOCA Mimicker. To our knowledge, this is the first study to evaluate STE-MINOCA according to the AHA guidelines. Compared to STEMI-Obstruction, unadjusted 5-year survival was significantly lower in AHA STE-MINOCA Mimicker but similar in AHA STE-MINOCA.

Among the 3 sites in the Midwest STEMI consortium, only one site had a standardized process to perform CMR for AHA STE-MINOCA cases. CMR substantially improved the diagnosis of the cause of AHA STE-MINOCA. At the site using CMR, 11% of AHA STE-MINOCA cases were classified as unknown etiology, compared to 43-56% at the other sites. These results are consistent with the Women’s Heart Attack Research Program (HARP), which demonstrated that CMR identified the probable mechanism of MINOCA in 74% of patients in a cohort almost entirely composed of NSTEMIs (96.5%) (24). Additional coronary intravascular imaging, such as optical coherence tomography (OCT), led to a diagnosis in 85% of MINOCA cases (24). A standardized diagnostic evaluation of STE-MINOCA that includes the use of CMR, intravascular imaging, and in selected cases, functional coronary angiography with coronary vasospasm provocation and coronary flow reserve assessment to determine the underlying mechanisms of MINOCA to further risk stratify patients and treat appropriately is needed (24–27).

There is growing evidence for the benefits of statins and renin-angiotensin system blockers for secondary prevention in MINOCA patients (28,29). Choo et al. demonstrated that the nonuse of renin-angiotensin system blockers and statin was associated with a 2-fold higher mortality risk in MINOCA patients (12). Notably, we documented that STE-MINOCA patients were less likely to be prescribed aspirin, beta-blockers, ACEI/ARB, statin, and P2Y12 inhibitors at discharge compared to STEMI-Obstruction. Others have also noted this treatment disparity in cohorts predominantly composed of MINOCA without STE (30,31). The higher mortality in STE-MINOCA emphasizes the need for secondary prevention to improve outcomes in this important patient subset.

### Limitations

This study has limitations common to observational studies, including unmeasured confounders and event adjudication. Angiographic images were not evaluated by a core lab; only cases identified as no culprit at the time of angiography and suspicious for STE-MINOCA were reviewed by two experts to classify based on current AHA definitions. The use of CMR in the evaluation of MINOCA was not standardized among the sites.

## Conclusions

In this large multi-center prospective cohort of consecutive STEMIs, nearly 5% of patients presented with STE-MINOCA. At five years, all-cause mortality approached 20% among patients with STE-MINOCA. Despite the lower traditional cardiovascular risk profile, STE-MINOCA patients were at 40% higher risk of all-cause mortality compared to STEMI-Obstruction. Compared to STEMI-Obstruction, all-cause mortality was higher in AHA STE-MINOCA Mimicker but similar in AHA STE-MINOCA. STE-MINOCA patients had a lower frequency of cardiac medications prescribed at hospital discharge, emphasizing the need to diagnose the underlying mechanism and, thereby implement effective treatments.

## Data Availability

Data openly available in a public repository that issues datasets with DOIs

## ABBREVIATIONS

MINOCA: Myocardial infarction with non-obstructive coronary arteries
AMI: Acute myocardial infarction
STEMI: ST-segment elevation myocardial infarction
STE-MINOCA: ST-segment elevation myocardial infarction with non-obstructive coronary arteries
CAD: Coronary artery disease
CVD: Cardiovascular disease
STEMI-Obstruction: ST-segment elevation myocardial infarction with coronary artery obstruction
AHA: American Heart Association
ESC: European Society of Cardiology
NSTEMI: Non-ST-segment elevation myocardial infarction
CMR: Cardiac magnetic resonance

## FIGURE LEGENDS

**Supplemental Figure 1.**
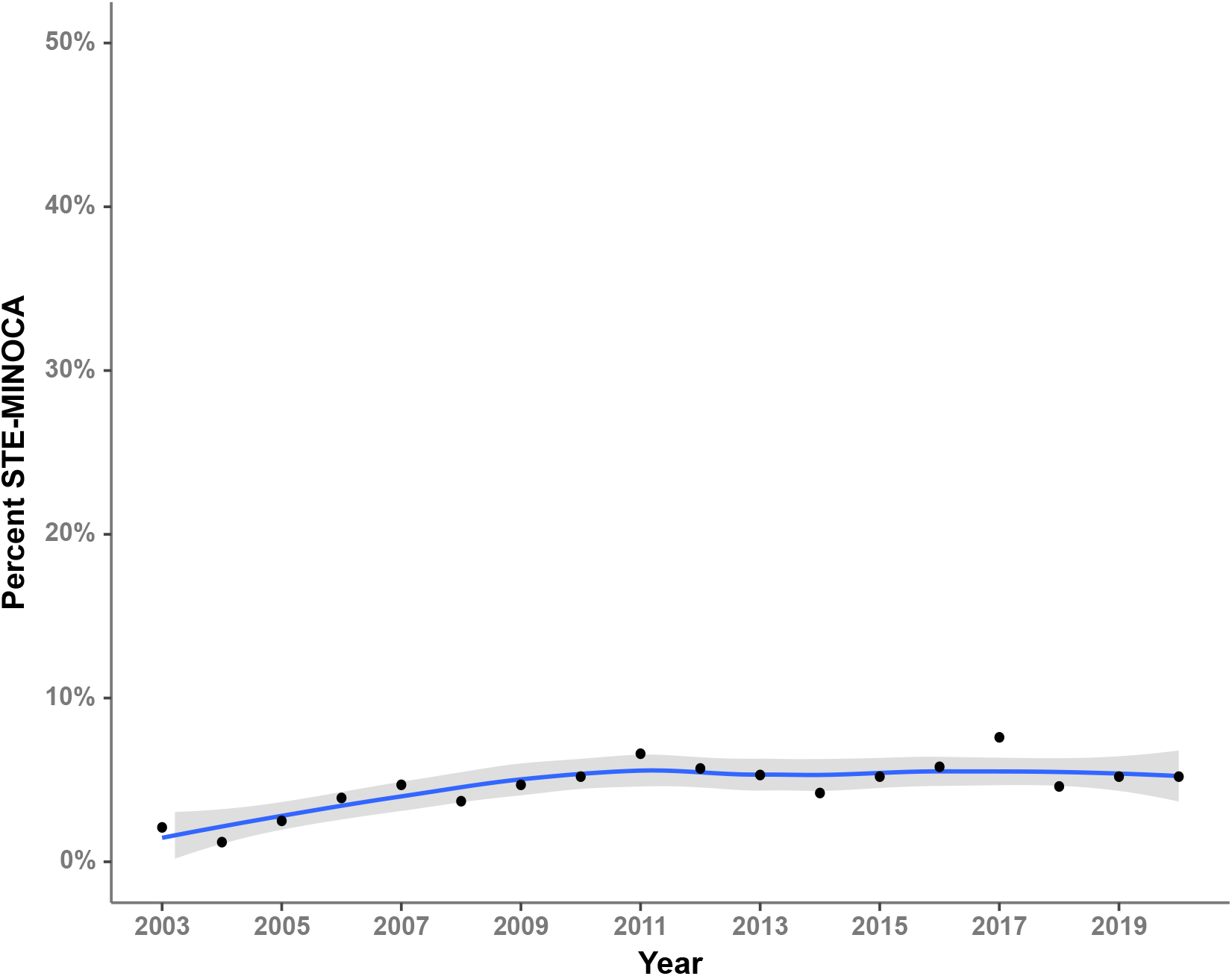
Distribution of the frequency to STE-MINOCA during the enrollment period by a year.

**Supplemental Figure 2.**
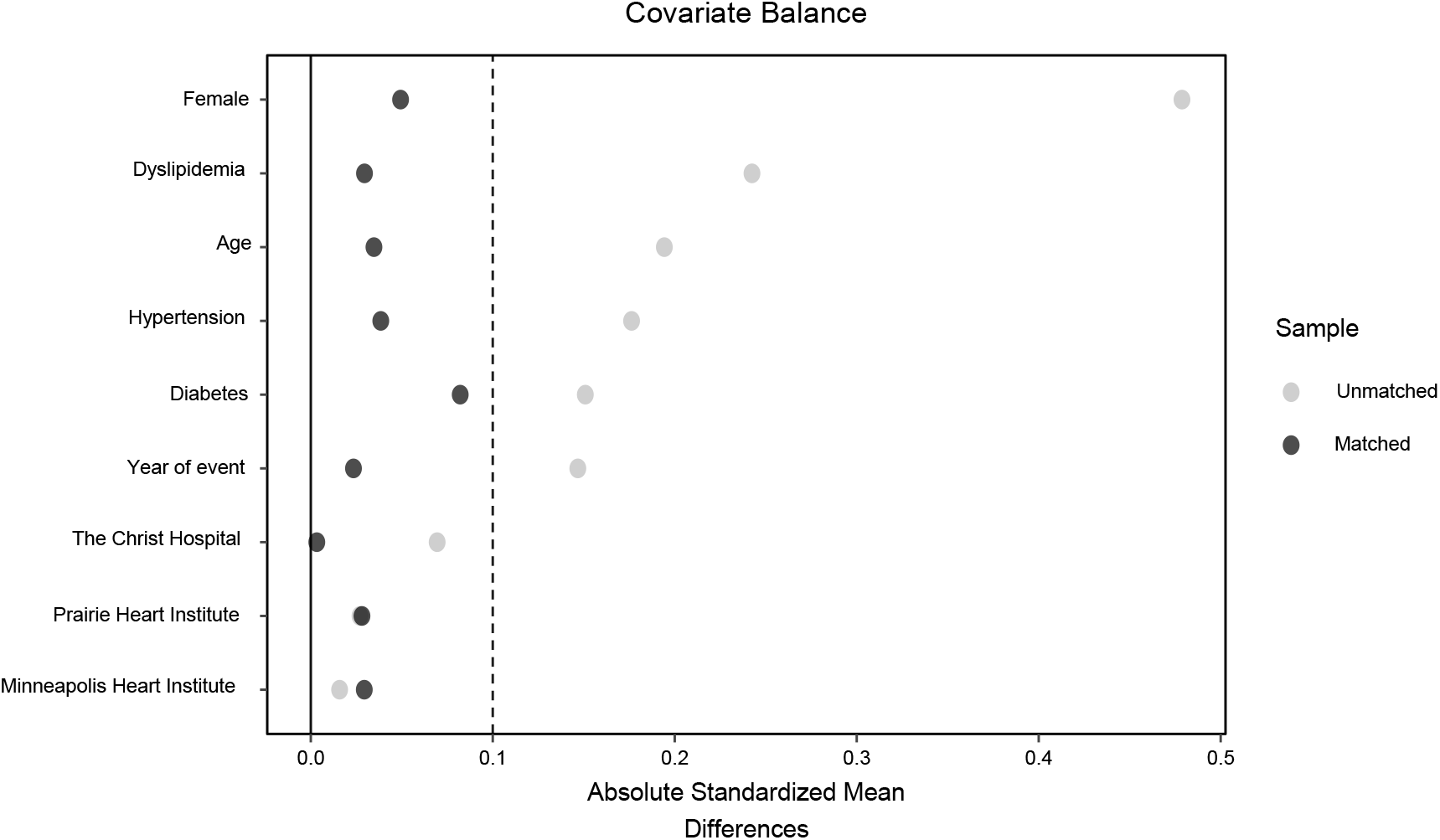
Covariate Balance Plot for Propensity Matched Cohort Analysis of 5-Year Survival in STE-MINOCA in Comparison with STEMI-Obstruction.

**Central Figure.**
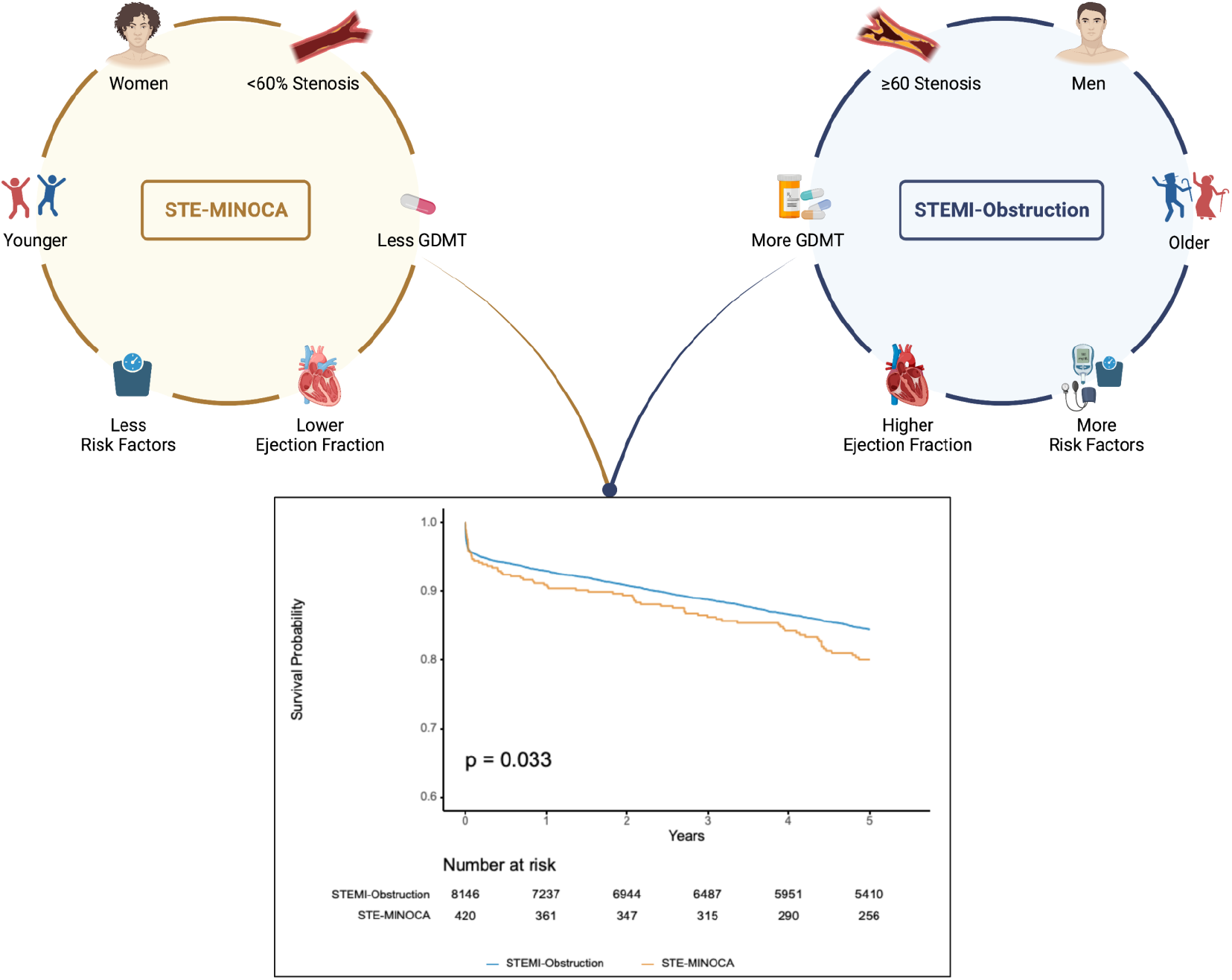
Differences in Risk Profile and 5-Year Mortality in STEMI-Obstruction Compared to STE-MINOCA.

**Supplemental Table 1.**
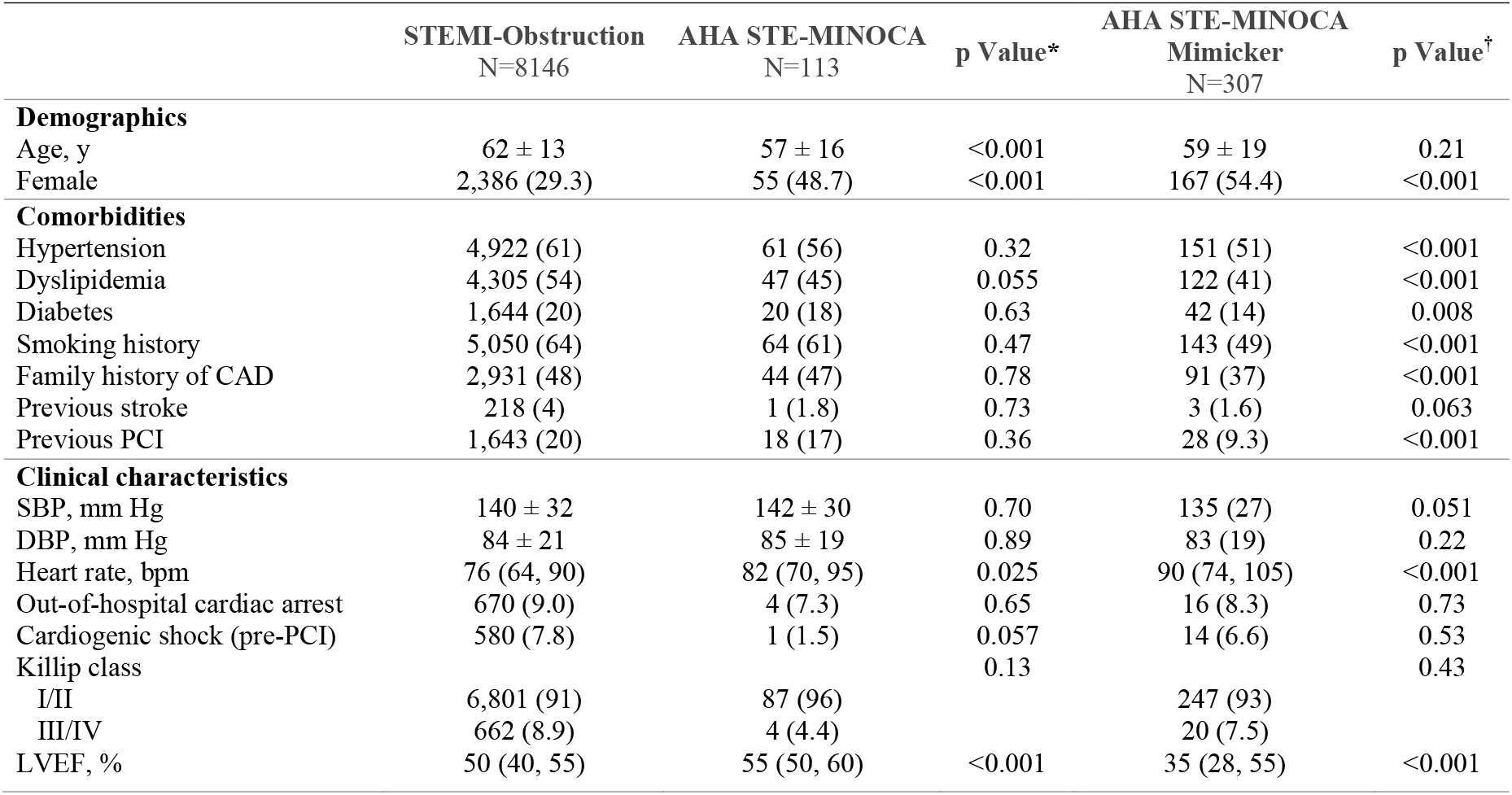
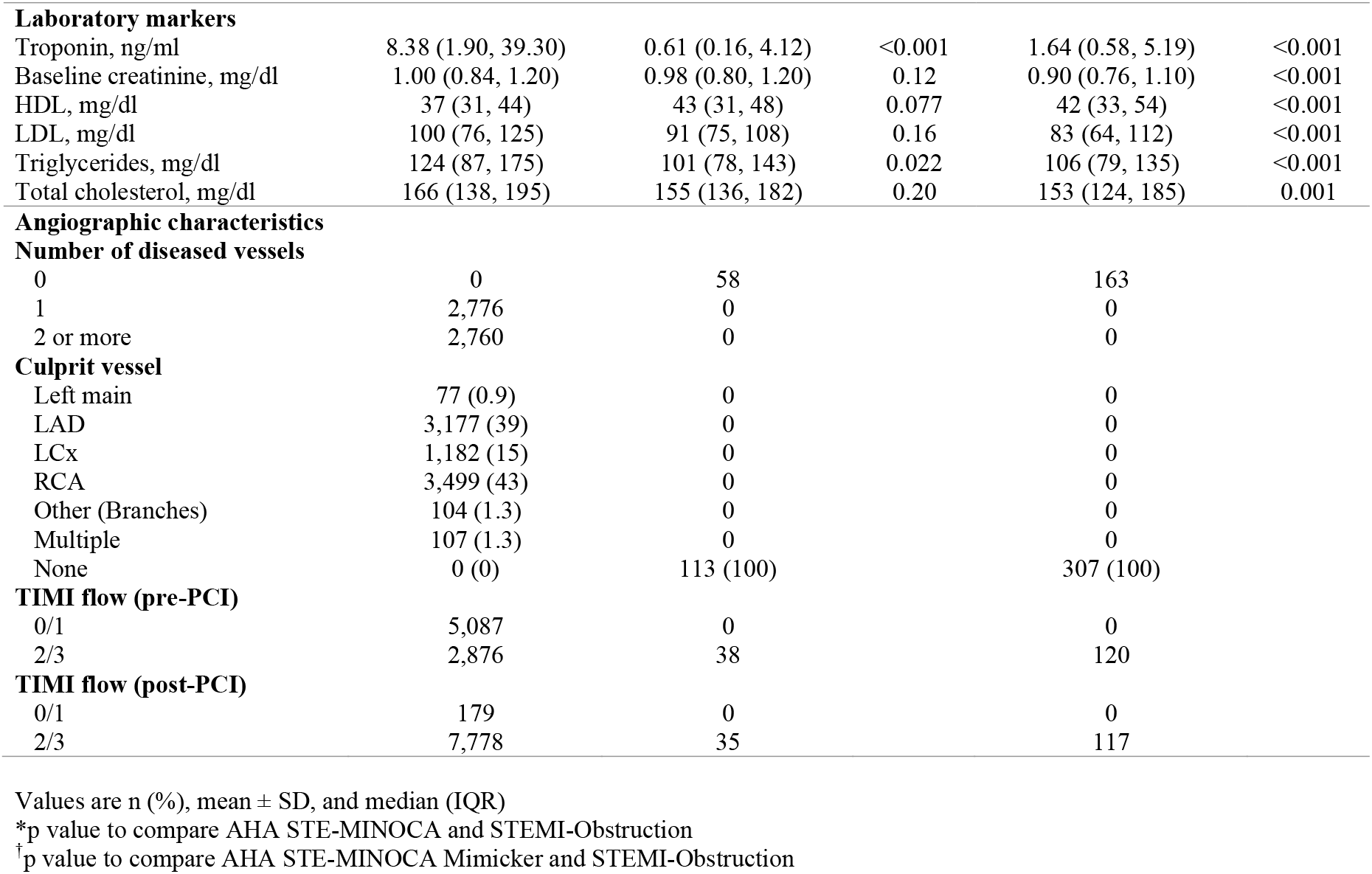
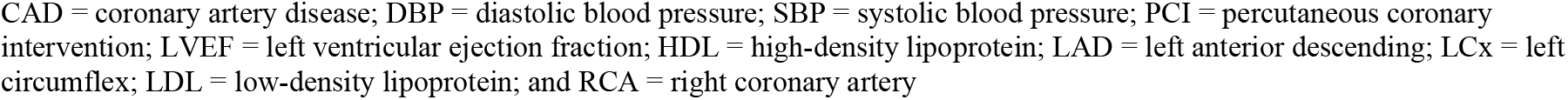
Demographics and Clinical Characteristics in STEMI-Obstruction compared to AHA STE-MINOCA or AHA STE-MINOCA Mimicker.

|  | STEMI-Obstruction<br>N=8146 | AHA STE-MINOCA<br>N=113 | p Value* | AHA STE-MINOCA<br>Mimicker<br>N=307 | p Value<sup>†</sup> |
| --- | --- | --- | --- | --- | --- |
| **Demographics** |  |  |  |  |  |
| Age, y | 62 ± 13 | 57 ± 16 | <0.001 | 59 ± 19 | 0.21 |
| Female | 2,386 (29.3) | 55 (48.7) | <0.001 | 167 (54.4) | <0.001 |
| **Comorbidities** |  |  |  |  |  |
| Hypertension | 4,922 (61) | 61 (56) | 0.32 | 151 (51) | <0.001 |
| Dyslipidemia | 4,305 (54) | 47 (45) | 0.055 | 122 (41) | <0.001 |
| Diabetes | 1,644 (20) | 20 (18) | 0.63 | 42 (14) | 0.008 |
| Smoking history | 5,050 (64) | 64 (61) | 0.47 | 143 (49) | <0.001 |
| Family history of CAD | 2,931 (48) | 44 (47) | 0.78 | 91 (37) | <0.001 |
| Previous stroke | 218 (4) | 1 (1.8) | 0.73 | 3 (1.6) | 0.063 |
| Previous PCI | 1,643 (20) | 18 (17) | 0.36 | 28 (9.3) | <0.001 |
| **Clinical characteristics** |  |  |  |  |  |
| SBP, mm Hg | 140 ± 32 | 142 ± 30 | 0.70 | 135 (27) | 0.051 |
| DBP, mm Hg | 84 ± 21 | 85 ± 19 | 0.89 | 83 (19) | 0.22 |
| Heart rate, bpm | 76 (64, 90) | 82 (70, 95) | 0.025 | 90 (74, 105) | <0.001 |
| Out-of-hospital cardiac arrest | 670 (9.0) | 4 (7.3) | 0.65 | 16 (8.3) | 0.73 |
| Cardiogenic shock (pre-PCI) | 580 (7.8) | 1 (1.5) | 0.057 | 14 (6.6) | 0.53 |
| Killip class |  |  | 0.13 |  | 0.43 |
| I/II | 6,801 (91) | 87 (96) |  | 247 (93) |  |
| III/IV | 662 (8.9) | 4 (4.4) |  | 20 (7.5) |  |
| LVEF, % | 50 (40, 55) | 55 (50, 60) | <0.001 | 35 (28, 55) | <0.001 |
| **Laboratory markers** |  |  |  |  |  |
| Troponin, ng/ml | 8.38 (1.90, 39.30) | 0.61 (0.16, 4.12) | <0.001 | 1.64 (0.58, 5.19) | <0.001 |
| Baseline creatinine, mg/dl | 1.00 (0.84, 1.20) | 0.98 (0.80, 1.20) | 0.12 | 0.90 (0.76, 1.10) | <0.001 |
| HDL, mg/dl | 37 (31, 44) | 43 (31, 48) | 0.077 | 42 (33, 54) | <0.001 |
| LDL, mg/dl | 100 (76, 125) | 91 (75, 108) | 0.16 | 83 (64, 112) | <0.001 |
| Triglycerides, mg/dl | 124 (87, 175) | 101 (78, 143) | 0.022 | 106 (79, 135) | <0.001 |
| Total cholesterol, mg/dl | 166 (138, 195) | 155 (136, 182) | 0.20 | 153 (124, 185) | 0.001 |
| **Angiographic characteristics** |  |  |  |  |  |
| **Number of diseased vessels** |  |  |  |  |  |
| 0 | 0 | 58 |  | 163 |  |
| 1 | 2,776 | 0 |  | 0 |  |
| 2 or more | 2,760 | 0 |  | 0 |  |
| **Culprit vessel** |  |  |  |  |  |
| Left main | 77 (0.9) | 0 |  | 0 |  |
| LAD | 3,177 (39) | 0 |  | 0 |  |
| LCx | 1,182 (15) | 0 |  | 0 |  |
| RCA | 3,499 (43) | 0 |  | 0 |  |
| Other (Branches) | 104 (1.3) | 0 |  | 0 |  |
| Multiple | 107 (1.3) | 0 |  | 0 |  |
| None | 0 (0) | 113 (100) |  | 307 (100) |  |
| **TIMI flow (pre-PCI)** |  |  |  |  |  |
| 0/1 | 5,087 | 0 |  | 0 |  |
| 2/3 | 2,876 | 38 |  | 120 |  |
| **TIMI flow (post-PCI)** |  |  |  |  |  |
| 0/1 | 179 | 0 |  | 0 |  |
| 2/3 | 7,778 | 35 |  | 117 |  |

Values are n (%), mean ± SD, and median (IQR)
\*p value to compare AHA STE-MINOCA and STEMI-Obstruction
<sup>†</sup>p value to compare AHA STE-MINOCA Mimicker and STEMI-Obstruction
CAD = coronary artery disease; DBP = diastolic blood pressure; SBP = systolic blood pressure; PCI = percutaneous coronary intervention; LVEF = left ventricular ejection fraction; HDL = high-density lipoprotein; LAD = left anterior descending; LCx = left circumflex; LDL = low-density lipoprotein; and RCA = right coronary artery

**Supplemental Table 2.**
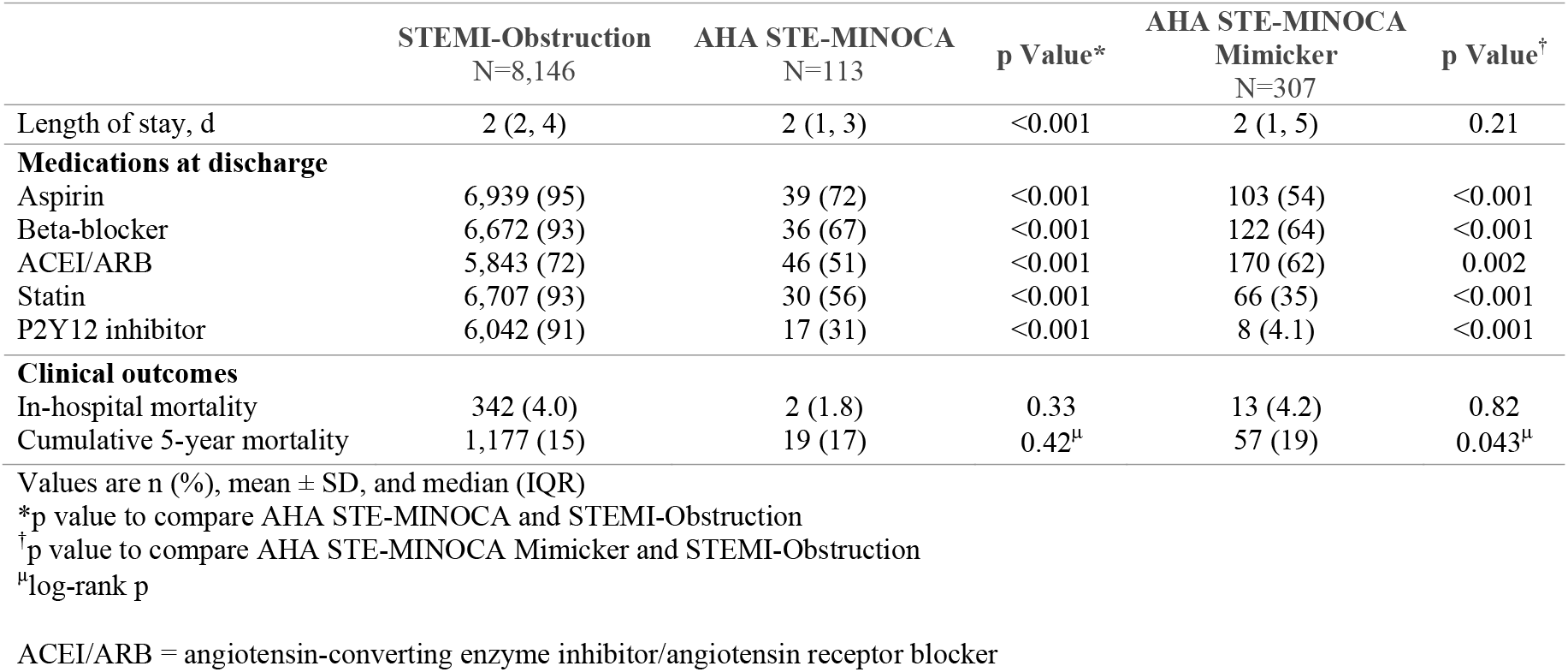
Medications and Outcomes in STEMI-Obstruction compared to AHA STE-MINOCA or AHA STE-MINOCA Mimicker.

